# Serum Potassium Monitoring using AI-enabled Smart Watch Electrocardiograms

**DOI:** 10.1101/2024.05.08.24307064

**Authors:** I-Min Chiu, Po-Jung Wu, Huan Zhang, J. Weston Hughes, Albert J Rogers, Laleh Jalilian, Marco Perez, Chun-Hung Richard Lin, Chien-Te Lee, James Zou, David Ouyang

**Affiliations:** Department of Cardiology, Smidt Heart Institute, Cedars-Sinai Medical Center, Los Angeles, CA, USA; Department of Emergency Medicine, Chang Gung Memorial Hospital Kaohsiung Branch, Kaohsiung, Taiwan; Department of Computer Science and Engineering, National Sun Yat-sen University, Kaohsiung, Taiwan; Division of Nephrology, Department of Internal Medicine, Kaohsiung Chang Gung Memorial Hospital, Kaohsiung, Taiwan; Department of Computer Science, Stanford University, Palo Alto, CA, USA; Department of Medicine and Cardiovascular Institute, Stanford University School of Medicine, Stanford, CA, USA; Department of Anesthesiology and Perioperative Medicine, University of California Los Angeles, Los Angeles, CA; Division of Nephrology, Department of Internal Medicine, Kaohsiung Chang Gung Memorial Hospital and Kaohsiung Municipal Feng-Shan Hospital, Kaohsiung, Taiwan; Department of Biomedical Data Science, Stanford University, Palo Alto, CA, USA

**Keywords:** Potassium Monitoring, Hyperkalemia, Artificial Intelligence, Smart Watch, Apple Watch, Electrocardiograms

## Abstract

**Background:** Hyperkalemia poses a significant risk of sudden cardiac death, especially for those with end-stage renal diseases (ESRD). Smartwatches with ECG capabilities offer a promising solution for continuous, non-invasive monitoring using AI.

**Objectives:** To develop an AI-ECG algorithm to predict serum potassium level in ESRD patient with smartwatch generated ECG waveforms.

**Methods:** A cohort of 152,508 patients with 293,557 ECGs paired serum potassium levels obtained within one hour at Cedars Sinai Medical Center (CSMC) was used to train an AI-ECG model (‘Kardio-Net’) to predict serum potassium level. The model was further fine-tuned on 4,337 ECGs from 1,463 patients with ESRD using inputs from 12-lead and single-lead ECGs. Kardio-Net was evaluated in held-out test cohorts from CSMC and Stanford Healthcare (SHC) as well as a prospective international cohort of 40 ESRD patients with smartwatch ECGs at Chang Gung Memorial Hospital (CGMH).

**Results:** The Kardio-Net, when applied to 12-lead ECGs, identified severe hyperkalemia with an AUC of 0.852 and a mean absolute error (MAE) of 0.527 mEq/L. In external validation at SHC, the model achieved an AUC of 0.849 and an MAE of 0.599 mEq/L. For single-lead ECGs, Kardio-Net detected hyperkalemia with an AUC of 0.876 and had an MAE of 0.575 mEq/L in the CSMC test cohort. Using prospectively obtained smartwatch data, the AUC was 0.831, with an MAE of 0.580 mEq/L.

**Conclusions:** We validate a deep learning model to predict serum potassium levels from both 12-lead ECGs and single-lead smartwatch data, demonstrating its utility for remote monitoring of hyperkalemia.

**Condensed Abstract:** Hyperkalemia significantly increases the risk of sudden cardiac death in end-stage renal disease (ESRD) patients. We developed ‘Kardio-Net,’ an AI-driven ECG model, using data from 152,508 patients at Cedars Sinai Medical Center, and refined it with ECGs from 1,463 ESRD patients using inputs from 12-lead and single-lead ECGs. This model facilitates continuous and non-invasive potassium monitoring, leveraging both traditional and smartwatch-generated ECGs. Tested across various cohorts, including a prospective smartwatch group, Kardio-Net achieved an AUC range of 0.807 to 0.876, demonstrating its effectiveness for real-time hyperkalemia monitoring.

## Introduction

Hyperkalemia, or elevated serum potassium, presents a severe and potentially life-threatening risk, in particular for patients with acute kidney injury or chronic kidney disease. Patients with end-stage renal disease (ESRD)^1^ are at increased risk for sudden cardiac death due to hyperkalemia^2–4^ and require vigilant electrolyte monitoring for hemodialysis^3^. Meanwhile, phlebotomy for serum potassium monitoring is resource-intensive, invasive, and limited in ability to monitor in real time. A noninvasive approach for potassium monitoring could greatly improve the detection of potential lethal electrolyte imbalances. Remote monitoring with smartwatch devices presents a potential solution by offering the capability for remote health monitoring.

There are well known changes in electrocardiogram (ECG) with changes in serum potassium, including the peaking of T waves, QRS prolongation, and PR shortening^5^. Despite recognizable changes, hyperkalemia associated changes are subtle, leading to low sensitivity of physician readers^6^. Recent advancements in artificial intelligence (AI), especially deep learning (DL), have shown significant promise in enhancing the analysis and interpretation of ECG signals^7–13^. Research in the past few years has been instrumental in improving the sensitivity for detecting hyperkalemia, demonstrating the potential of AI to augment clinical decision-making in identifying this dangerous electrolyte imbalance^14–17^. Recent work have shown the ability for AI-ECG applications originally developed using 12-lead ECGs^18^ to be optimized for single lead smartwatch ECGs^13^.

We hypothesized an AI-enhanced ECG model (Kardio-Net) can accurately identify hyperkalemia, including in patients with ERSD using ECG data collected from a smartwatch. To test our hypothesis, we develop, validate, and test a model for predicting potassium levels across three international institutions using both 12-lead and smartwatch ECGs in ESRD patients.

## Method

### Patient identification and data sources

A primary cohort of patients with paired ECGs and serum potassium levels was curated from the CSMC between 2008 to 2022. A total of 153,971 adult patients had at least one 12-lead ECG and serum potassium test within one hour, leading to 297,914 twelve-lead ECGs and corresponding potassium values. ECGs were mapped to the temporally closest potassium test. Given an interest in assessing performance in the ESRD population, all recordings from non ESRD patients (n = 152,508) were used for initial model training, and the model was then finetuned on ESRD patients alone (n = 1,463). The cohorts were randomly split at the patient level into 80% for training set, 10% for validation, and 10% reserved as a held-out test set.

For external validation, we identified patient cohorts from two other healthcare systems. We identified 7,586 ECGs among 3,107 ESRD patients at Stanford HealthCare (SHC) from 8/2005 to 6/2018 (Figure 1). Additionally, A prospective cohort of patients with ESRD at Chang Gung Memorial Hospital (CGMH) were enrolling from October 2022 to July 2023 with serial smartwatch based ECG testing prior to hemodialysis. Prior to participation, written consent was secured from each individual. This study was approved by the institutional review boards of the CSMC, SHC, and CGMH. Patients’ informed consent was waived at CSMC and SHC due to the retrospective nature of the study using de-identified ECG and electronic health record data.

**Figure 1:**
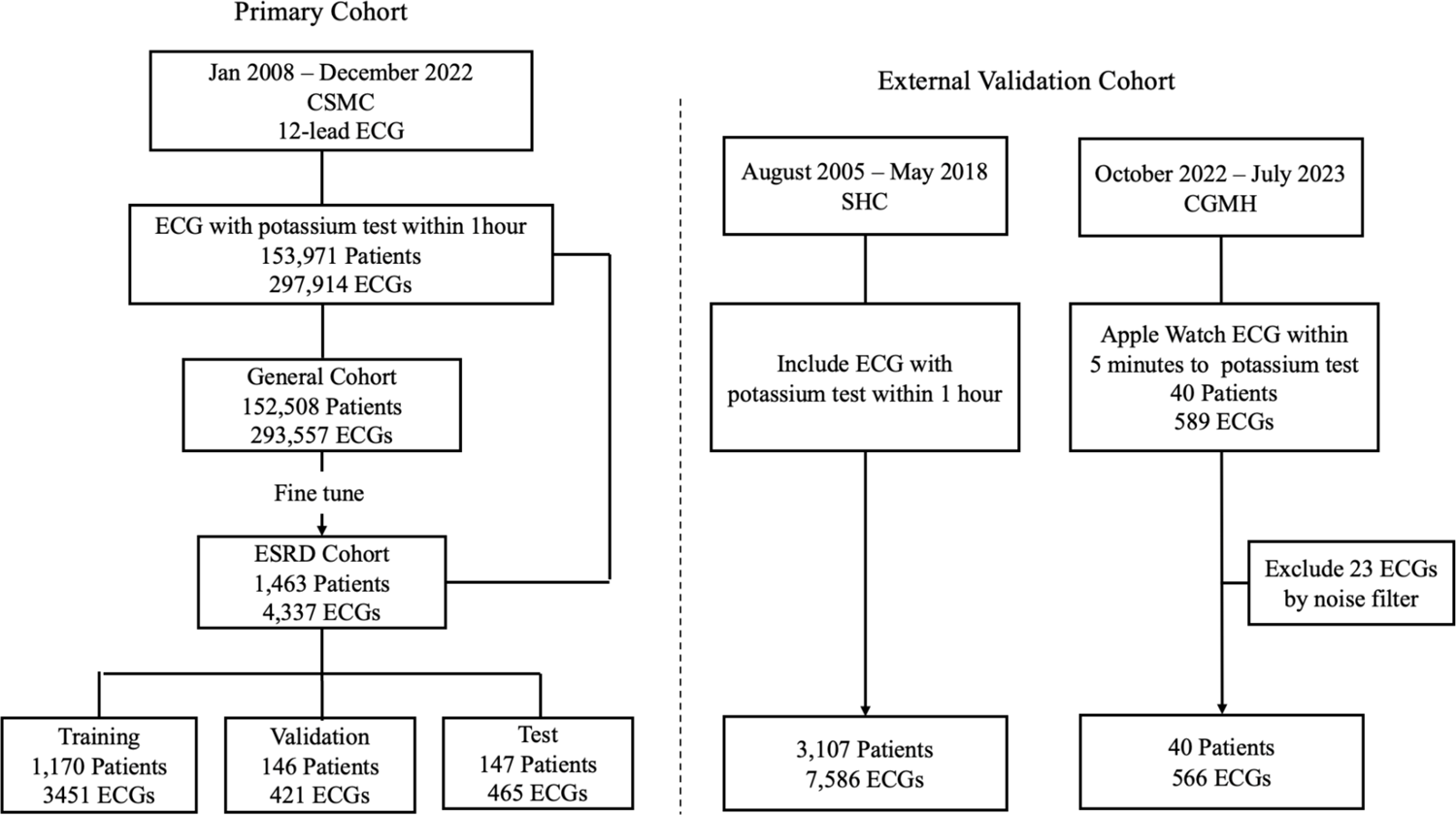
Selection of study subjects in the primary and external validation cohorts.

### Model Development

We developed a convolutional neural network (CNN) designed for ECG interpretation, Kardio-Net, with the capability to integrate digitized waveform for predicting serum potassium levels. All ECGs obtained from CSMC were recorded using a GE machine, adhering to the standard 10-second, 12-lead ECG protocol. These ECG waveform data were captured at a sampling rate of 500 Hz and represented as 10-second, 12×5000 matrices of amplitude values. In external validation at SHC, ECGs were stored using the Phillips TraceMaster system with the same sampling rate with recorded also as 12×5000 matrices and were independently used as input examples for external validation.

Kardio-Net was trained on PyTorch starting from random initialization. Training utilized a mean square error loss for up to 100 epochs, employing an ADAM optimizer with a starting learning rate of 1e-2. Early stopping based on minimizing validation cohorts mean absolute error (MAE). For the fine-tuning in ESRD patients, weights from the initial model were used as the starting point and trained using the same loss function and optimizer with a learning rate of 1e-4. To ensure compatibility with the smartwatch ECG data, we trained the Kardio-Net by utilized lead I from the 12-lead ECGs, as it most closely resembles the ECG vector of smartwatch recordings. For Kardio-Net inference on ECG waveforms smartwatch, we split the waveform data into consecutive 5-second intervals. During the inference, a 30-second ECG waveform was split at 1-second intervals, yielding 26 5-second waveform data inputs. These were then averaged to formulate the final prediction.

### Validation on smartwatch ECG

After consent and enrollment of the patients, digital ECG recordings were captured by the Apple Watch ECG within 5 minutes before their routine serum potassium tests prior to hemodialysis. The smartwatch ECG waveforms, initially recorded at a sampling rate of 512 Hz, were resized to 500 Hz to match the sampling rate of 12 lead ECGs. The digitized ECG waveforms stored on the hospital server via the Apple Developer HealthKit API for subsequent analysis^20^. This process yielded a validation dataset of 589 ECGs waveforms paired with its corresponding potassium levels. Given that smartwatch waveforms can contain artifacts and baseline wandering distinct from 12-lead ECGs, we developed a noise ECG classifier to exclude excessively noisy waveforms before averaging the predictions for final analysis. 23 (3.9%) ECGs were excluded from the noise classifier. A domain adaptation technique was employed to facilitate the Kardio-Net’s application to Apple Watch ECG waveforms^19^. This adjustment aims to mitigate domain discrepancies, thereby enhancing Kardio-Net‘s performance on Apple Watch ECGs.

### Statistical analysis

All analyses were conducted on the held-out test dataset, external validation set, and smartwatch dataset, which were not used during the model’s training. The primary metric for assessing Kardio-Net’s performance in predicting serum potassium levels was the MAE. Additionally, the model’s ability to identify severe hyperkalemia, defined as potassium levels greater than 6.5 mEq/L, was evaluated using the area under the receiver operating characteristic curve (AUC). These metrics allow for a comprehensive assessment of the model’s classification accuracy and precision in detecting cases of hyperkalemia. For each metric, we calculated two-sided 95% confidence intervals using 1000 bootstrapped samples to ensure robust statistical inference. The modeling pipeline was implemented using Python (3.8) with PyTorch (2.0) as the deep learning framework. Signal processing and data analysis were facilitated by Python libraries such as SciPy (1.11), Scikit-learn (1.3.2), pandas (2.0.2), and matplotlib (3.7.1).

## Result

### Primary cohort characteristics

We identified a total of 153,971 patients at Cedars-Sinai Medical Center who had at least one pair of ECG and serum potassium measurement within 1 hour, resulting in 293,557 ECG recordings. 1,463 patients diagnosed with ESRD contributed 4,337 ECGs were selected as ESRD cohort, where other 293,557 ECGs from 152,508 patients formed the general cohort. The ESRD cohort was divided into a training set of 1,170 patients, a validation set of 146 patients, and a test set of 147 patients. The median age within the ESRD group was 62 years (interquartile range [IQR]: 50-72), with 57.8% (846 patients) being male. The median serum potassium level was 4.6 mEq/L (IQR: 4.1-5.2), with 190 ECGs (4.4%) indicating a potassium level higher than 6.5 mEq/L. Additional demographic and clinical characteristics are detailed in Table 1.

**Table 1.** Baseline Characteristics of Patients.

| Characteristic | CSMC |  |  | Apple watch | Stanford |
| --- | --- | --- | --- | --- | --- |
|  | Train | Val | Test |  |  |
| Number of patients | 1,170 | 146 | 147 | 40 | 3107 |
| Number of ECGs | 3,451 | 421 | 465 | 566 | 7586 |
| Sex (% Male) | 682 (58.3%) | 81 (55.5%) | 83 (56.5%) | 16 (40.0%) | 1820 (58.6%) |
| Age, median (IQR), y | 62 (50-72) | 61 (50-69) | 60 (50-72) | 65 (58-71) | 60 (49-70) |
| Race |  |  |  |  |  |
| Caucasian, n (%) | 621 (53.1%) | 76 (52.1%) | 73 (49.7%) | 0 | 1147 (36.9%) |
| Black, n (%) | 304 (26.0%) | 39 (26.7%) | 41 (27.9%) | 0 | 308 (9.9%) |
| Asian, n (%) | 140 (12.0%) | 19 (13.0%) | 24 (16.3%) | 40 (100%) | 542 (17.4%) |
| Others, n (%) | 105 (9.0%) | 12 (8.2%) | 9 (6.1%) | 0 | 1110 (35.7%) |
| Clinical |  |  |  |  |  |
| Atrial Fibrillation | 47 (4.0%) | 6 (4.1%) | 7 (4.8%) | 3 (7.5%) | 786 (25.3%) |
| Heart Failure | 222 (19.0%) | 16 (11.0%) | 26 (17.7%) | 4 (10.0%) | 1149 (37.0%) |
| CAD | 334 (28.5%) | 37 (25.3%) | 42 (28.6%) | 9 (22.5%) | 1087<br>(35.0%) |
| Hypertension | 534 (45.6%) | 63 (43.2%) | 79 (53.7%) | 26 (65.0%) | 2179<br>(70.1%) |
| Diabetes Mellitus | 401 (34.3%) | 45 (30.8%) | 56 (38.1%) | 15 (37.5%) | 1542<br>(48.7%) |
| Potassium (mEq/L) |  |  |  |  |  |
| median (IQR) | 4.6 (4.1-5.2) | 4.5 (4.0-5.2) | 4.5 (4.0-5.1) | 4.9 (4.4-5.3) | 4.5 (4.0-5.1) |
| >6.5, n (%) | 161 (4.7%) | 12 (2.9%) | 17 (3.7%) | 8 (1.5%) | 340 (4.5%) |

### Kardio-Net performance in the primary cohort

The initial deployment of our 12-lead Kardio-Net model, which was pre-trained on a general cohort, yielded an MAE of 0.593 mEq/L in the test set of ESRD cohort, with predictions concentrating around the 4.0 - 5.0 mEq/L level (Extended Figure 1). Upon fine-tuning this model with training set in ESRD cohort, there was a notable improvement in prediction accuracy, with the MAE reducing to 0.527 mEq/L. Kardio-Net achieved discrimination of hyperkalemia (potassium > 6.5 mEq/L) with an AUC of 0.852 (95% CI 0.745–0.956) (Figure 2).

**Figure 2:**
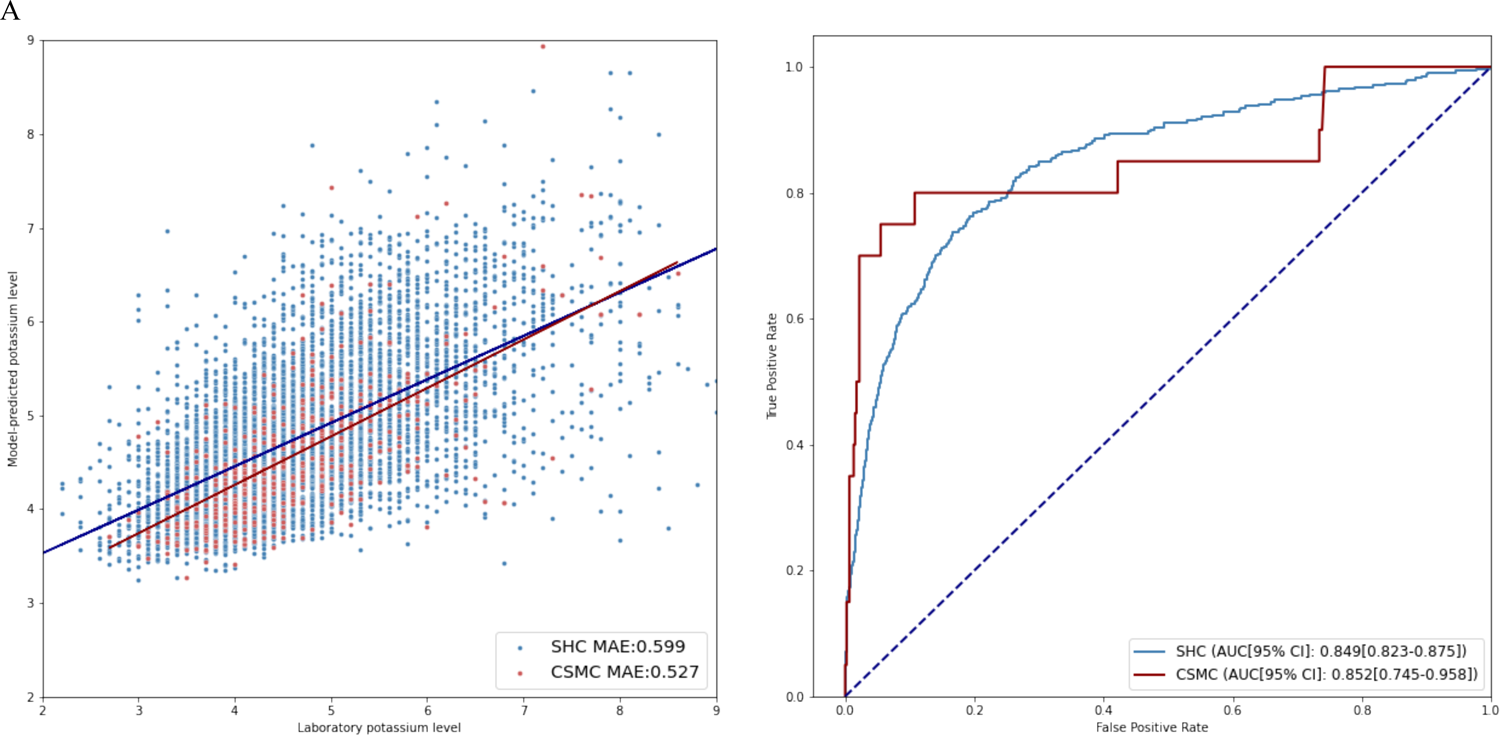
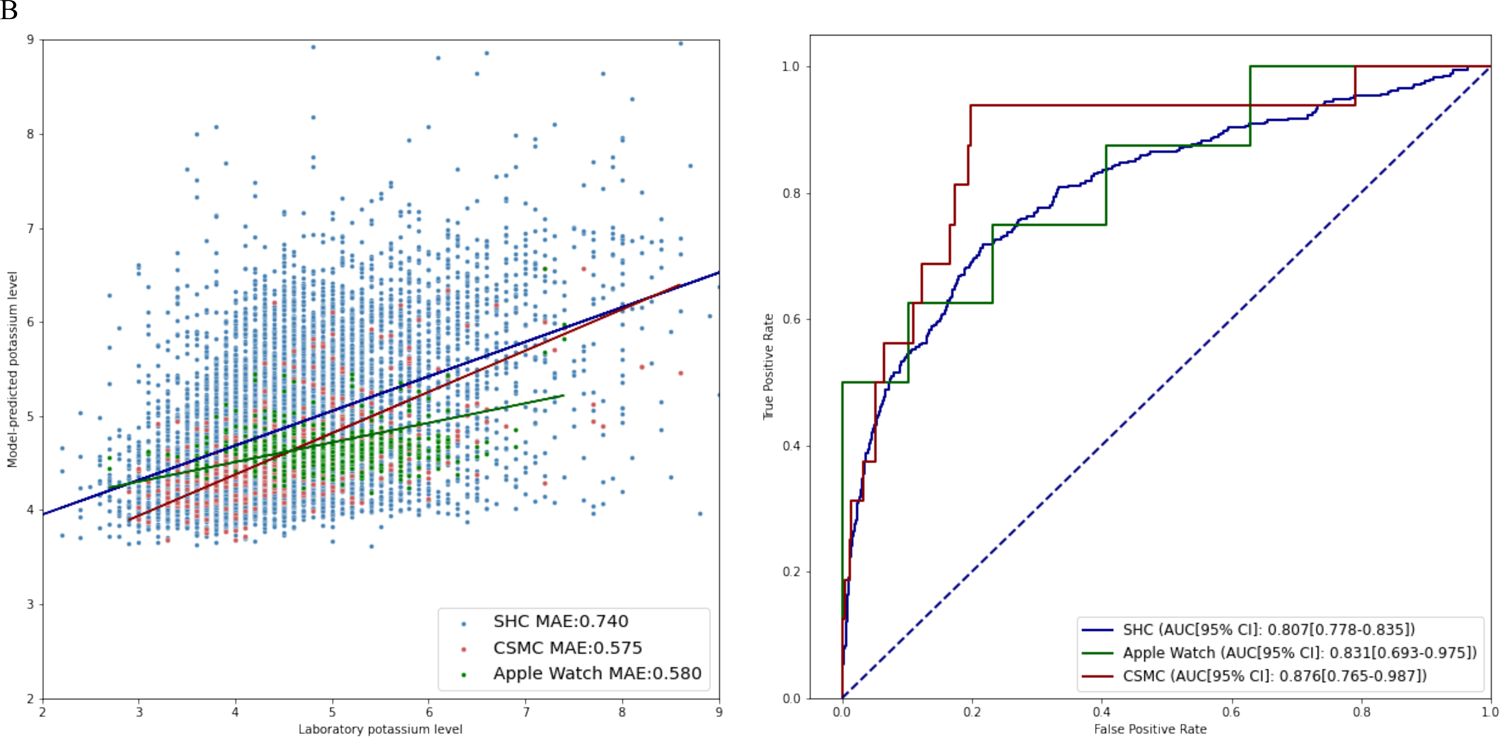
Deep learning model performance in internal and external validations. Figure 2A displays a scatter plot of serum potassium predictions from the 12-lead ECG model alongside the ROC curve for hyperkalemia recognition. Figure 2B presents the results from the single-lead ECG model.

To adapt to apple watch, we trained a single-lead ECG model using only lead I data through the same process. The model achieved an AUC of 0.876 (95% CI 0.765–0.987) on hyperkalemia detection and MAE of 0.575 mEq/L. Further evaluation of Kardio-Net’s performance across various patient subgroups, including older individuals and those with diabetes, hypertension, atrial fibrillation, heart failure, and coronary artery disease, was conducted. These groups are generally considered high-risk or exhibit ECG morphology changes. The model’s discrimination AUC for detecting hyperkalemia varied from 0.725 to 0.983 across these subgroups. Detailed subgroup results are shown in figure 3.

**Figure 3:**
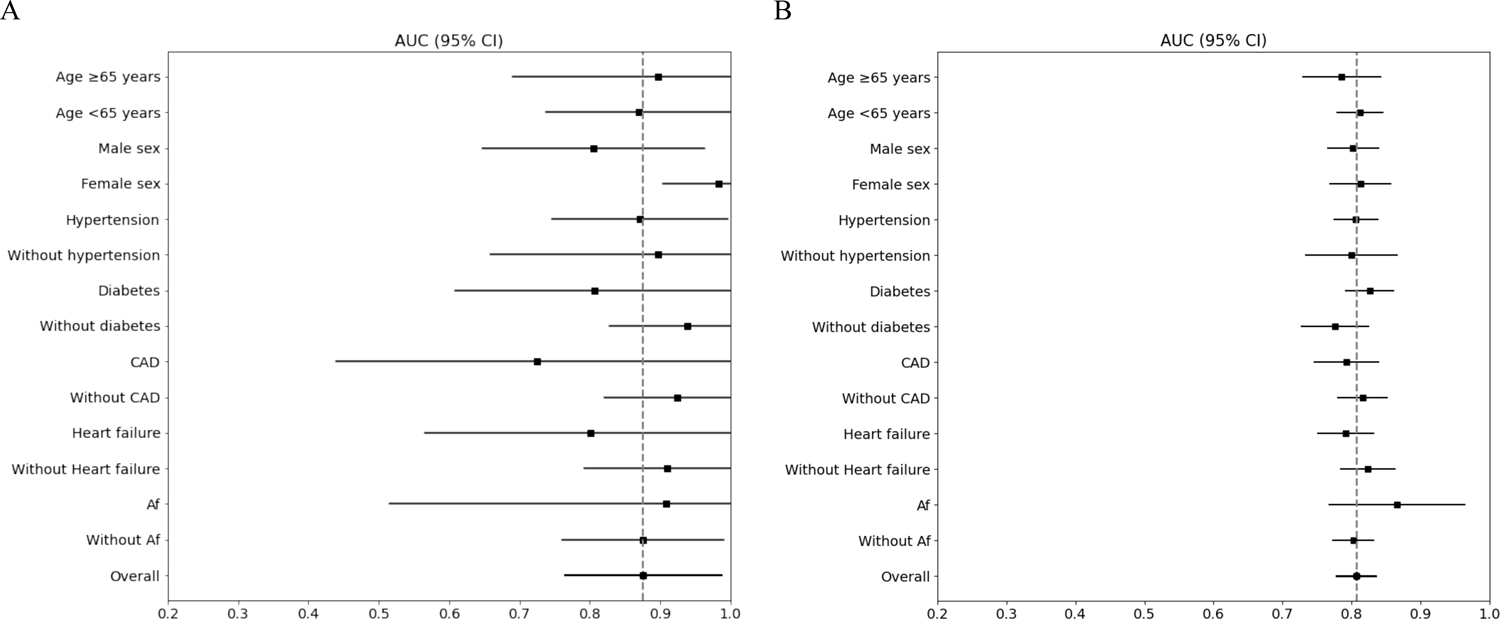
Subgroup analysis of hyperkalemia detection performance. Figure 3A illustrates the AUC for different subgroups within CSMC, and Figure 3B for SHC.

**Figure 4.**
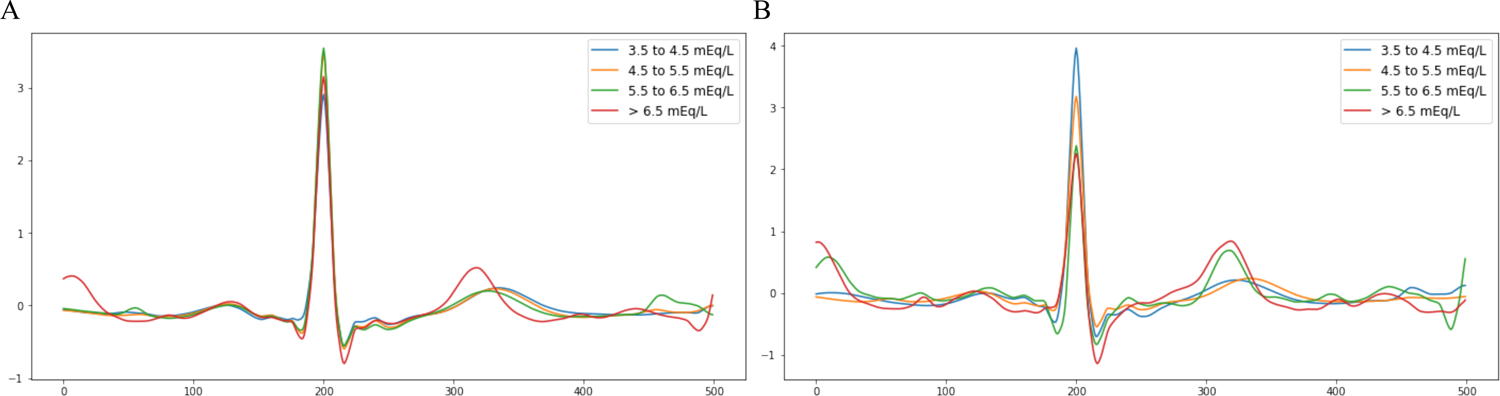
Correlated of ECG Waveforms with Serum Potassium Levels. Figure 4A shows mean ECG waveforms associated with measured serum potassium concentrations. The graph displays waveforms for potassium levels categorized into four ranges: 3.5 to 4.5 mEq/L (blue), 4.5 to 5.5 mEq/L (orange), 5.5 to 6.5 mEq/L (green), and >6.5 mEq/L (red), demonstrating the expected ECG changes corresponding to each potassium range. Figure 4B shows mean ECG waveforms corresponding to serum potassium level as predicted by Kardio-Net.

### Kardio-Net performance in the external validation dataset

The external validation cohort in SHC consisted of a total of 7,586 ECGs among 3,107 patients. The median age was 60 years (IQR: 49-70), with 58.6% (1,820 patients) being male. Atrial fibrillation is present in 25.3% of cases, heart failure in 37.0%, CAD in 35.0%, hypertension in 70.1%, and diabetes mellitus in 48.7%. The median serum potassium level was 4.5 mEq/L (IQR: 4.0-5.1), with 340 ECGs (4.5%) mapping to a severe hyperkalemia result. Other demographic and clinical characteristics are presented in Table 1.

In the external validation dataset, the performances of our 12-lead and single-lead Kardio-Net were consistent with the primary cohort. The 12-lead model demonstrated an AUC of 0.849 (95% CI 0.823-0.875) for hyperkalemia discrimination and achieved an MAE of 0.599 mEq/L. The single-lead model yielded an AUC of 0.807 (95% CI 0.778–0.835) and an MAE of 0.740 in predicting potassium levels. Further analysis in external validation was also conducted using the same subgroup to primary cohort. Kardio-Net’s discrimination AUC for detecting hyperkalemia varied from 0.780 to 0.860 across these subgroups with consistent performance.

### Validation in Smartwatch ECG Waveforms

In the prospective international cohort, 589 ECG waveforms were collected from 40 CGMH patients. The median age was 65 years ([IQR]: 58-71), with 16 male patients (40.0%). The median serum potassium level was 4.9 mEq/L (IQR: 4.4-5.3), with 8 ECGs (1.5%) associated with severe hyperkalemia. 23 (3.9%) ECGs were excluded for excessive noise and baseline wander. Single-lead Kardio-Net demonstrated an MAE of 0.580 mEq/L in predicting serum potassium base on smartwatch ECG. The model achieved an AUC of 0.831 (95% CI 0.693–0.975) for hyperkalemia detection using smartwatch ECG waveforms.

## Discussion

Hyperkalemia is a serious, potentially life-threatening electrolyte disturbance that significantly increases the risk for patients with ESRD, tripling their odds of mortality within one day when compared to those with normal potassium levels^21,22^. In this study, we developed a deep learning model, Kardio-Net, to estimate serum potassium levels using both standard 12-lead ECGs and single-lead ECGs including those from wearable devices like the Apple Watch. Across three geographically distinct institutions, our model was able to detect hyperkalemia in ESRD patients. These findings suggest potential for ongoing potassium monitoring in the periods between regular hemodialysis (Central Illustration).

Deep learning models have previously shown proficiency in detecting alterations in ECGs that indicate electrolyte imbalances^9,14,15,23,24^. Our research builds on existing studies that have used ECG data to predict potassium levels in patients with poor kidney function, who are particularly susceptible to the dangerous effects of hyperkalemia^4,21,25^. Earlier studies have suggested that certain ECG features can indicate potassium changes. However, these signs are often not as clear or accurate in patients with ESRD that their cardiac myocyte are less sensitive to changes in potassium, making the usual signs of high potassium less apparent^26–28^. Our study applying a deep learning approach that improves the precision of these predictions by training in a general patient and then finetune to enhanced performance specifically in the ESRD population. The transfer learning approach allows the model to retain what it learns from the much larger general population while adjusting its focus toward the ESRD population. This strategy is well documented in deep learning applications in biomedical research, where medical data are often sparse and more challenging to collect^29^.

Previous studies have shown the ability of DL models to detect hyperkalemia, but they also found that as the number of input leads decreased, the overall performance deteriorated^14^. Furthermore, the efficacy of algorithms specifically designed for wearable devices like the Apple Watch, where the signal-to-noise ratio is typically lower, had not been extensively tested^30^. Nevertheless, we adopted several approaches to enhance single-lead Kardio-Net performance as cited in previous literature, including a moving window average prediction from 5-s ECG^31^, and noise detection and removal^32^. These methods contributed the consistency of performance transit from 12-lead ECG to single lead ECG. This approach ensured the Kardio-Net’s prediction of serum potassium levels remained stable, with the MAE only slightly higher comparing 12-lead to single-lead ECGs.

### Study Limitations

Several limitations of the study merit consideration. First, the retrospective design of training data cohort may introduce selection bias of individuals who would receive 12-lead ECGs and serum electrolyte checks within 1 hour. This clinical setting might not reflect the variability found in broader outpatient settings or during daily activities where wearable technologies are used. However, we were able to evaluate the model’s performance on a prospectively collected cohort of patients immediately prior to hemodialysis. The prospective international cohort, which utilized smartwatch-generated waveforms from diverse racial groups, supports the generalizability of Kardio-Net. The use of real-world clinical data and the innovative application of deep learning techniques to a critical clinical biomarker substantially increase the relevance and potential impact of our findings in patient care management.

## Conclusion

Our research proposed a deep learning model, Kardio-Net, is capable of accurately predicting serum potassium levels in ESRD patients by analyzing both standard 12-lead ECGs and single-lead ECG data from wearable devices like the Apple Watch. The ability of this model to consistently identify hyperkalemia and its minimal increase in prediction error when moving from multi-lead to single-lead data supports its potential as a practical tool for continuous monitoring. Such technology could markedly enhance patient outcomes by facilitating the immediate detection of electrolyte imbalances, bridging the monitoring gap for patients undergoing regular hemodialysis.

### Perspectives

#### Competency in Medical Knowledge

The validation of an AI-ECG algorithm for real-time hyperkalemia monitoring via smartwatches offers a non-invasive, continuous potassium level assessment for high-risk patients.

#### Translational Outlook

Translating this AI-ECG innovation into clinical practice requires efficacy evaluations across diverse populations and integration with healthcare systems to better patient outcomes.

## Abbreviations

ESRD: End-Stage Renal Disease

ECG: Electrocardiogram

AI: Artificial Intelligence

DL: Deep Learning

SHC: Stanford HealthCare

CGMH: Chang Gung Memorial Hospital

CNN: Convolutional Neural Network

AUC: Area Under the receiver operating characteristic Curve

MAE: Mean Absolute Error

IQR: Interquartile Range

## Data Availability

The data are not publicly available due to restrictions.

---

**Central Illustration:**
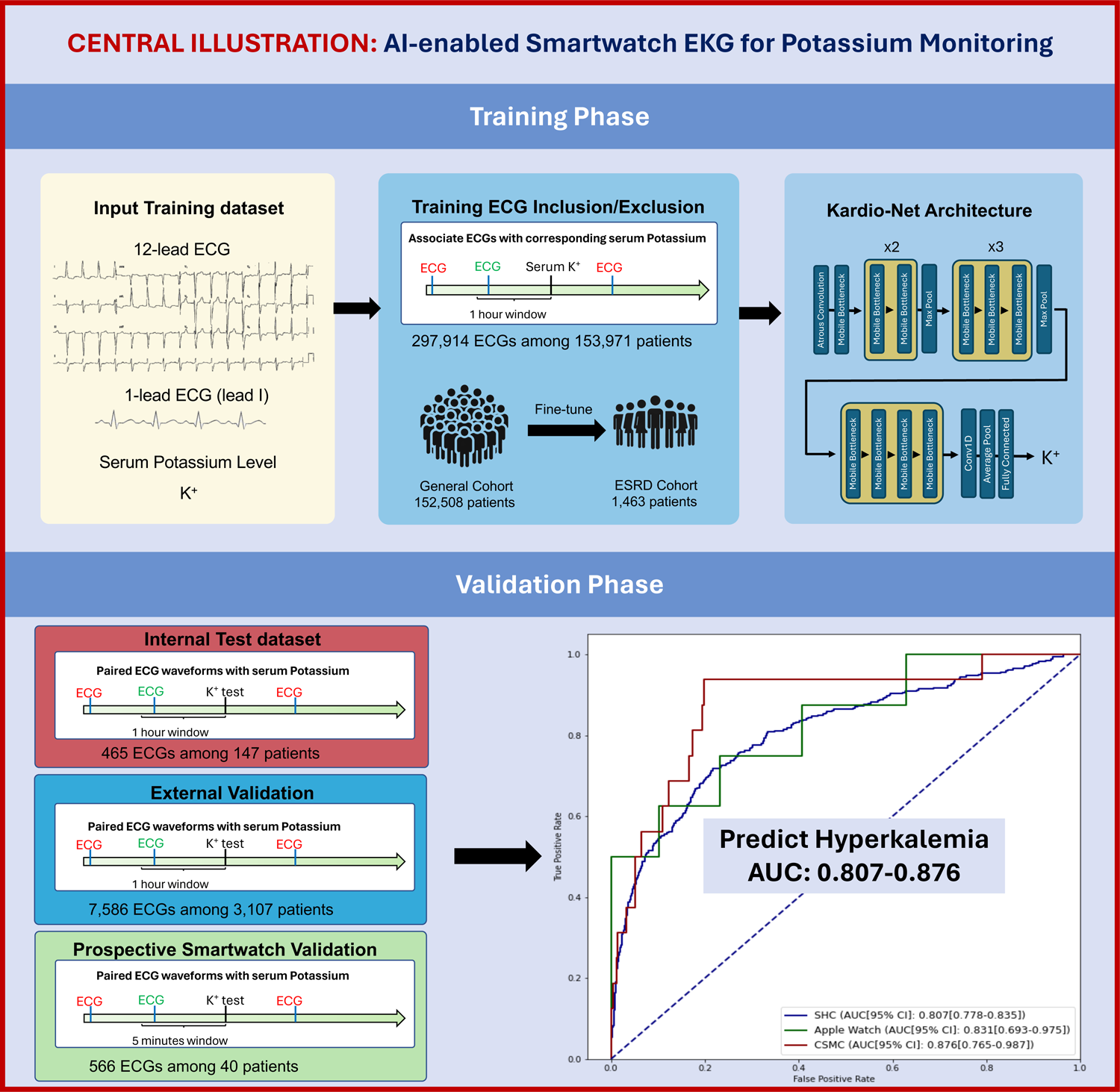
Kardio-Net is a convolutional neural network (CNN) designed to predict serum potassium levels in patients with end-stage renal disease (ESRD) using ECG data. Capable of processing both 12-lead and single-lead ECGs, this model adapts to traditional and smartwatch-based systems to provide continuous monitoring. Evaluated on retrospective and prospective cohorts, including smartwatch ECGs, Kardio-Net demonstrates robust performance with an AUC ranging from 0.807 to 0.876 in detecting hyperkalemia.

**Extended Figure 1:**
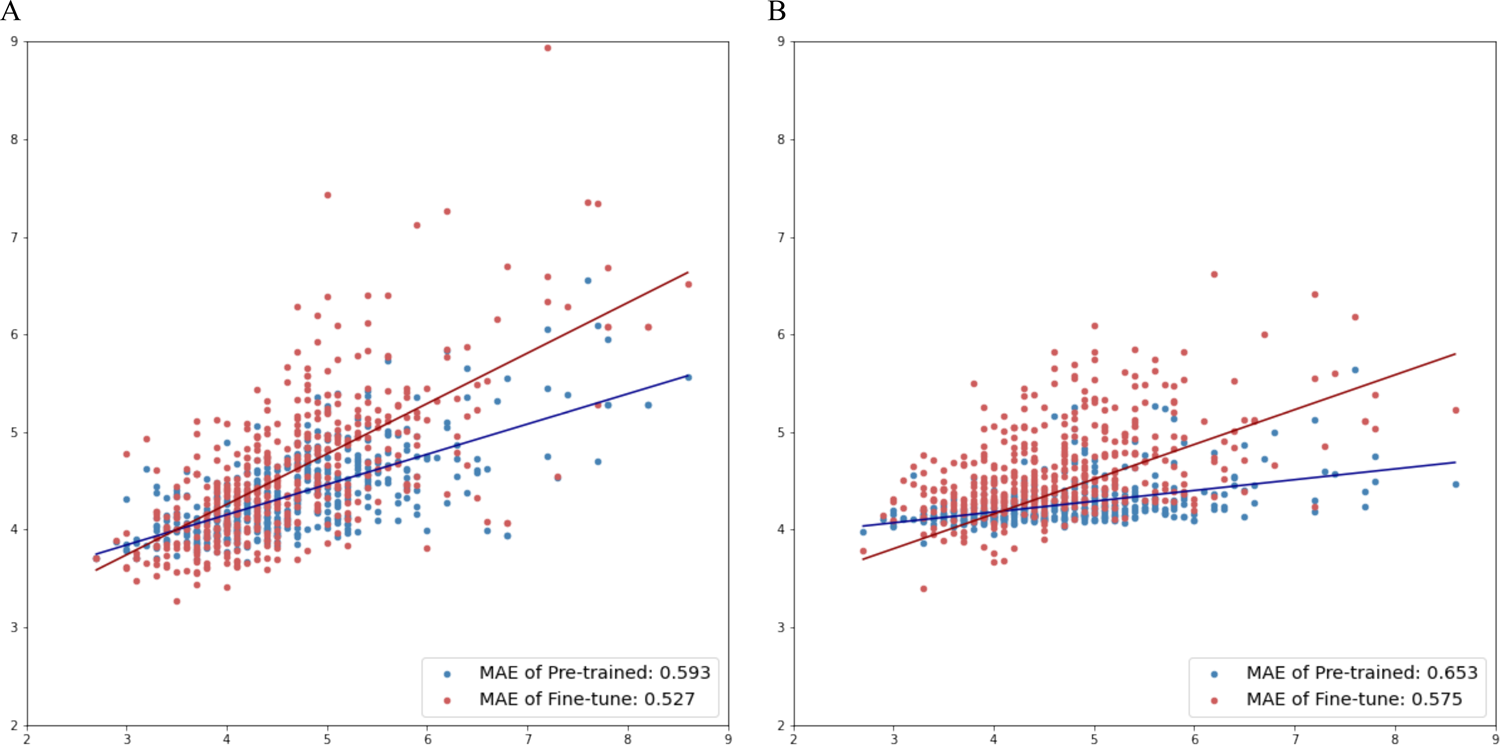
Scatter plots and MAE illustrating improved performance of the model in predicting serum potassium levels in the primary ESRD patient cohort after fine-tuning compared to the pre-training phase, using data from 12-lead (A) and single-lead (B) ECGs.

## Notes

### Competing Interest Statement

The authors have declared no competing interest.

### Funding Statement

I.M.C receive funding from National Science and Technology Council in Taiwan (111-2221-E-182A-008-); D.O and I.M.C receive funding from Apple and the NIH.

### Author Declarations

This study was approved by the institutional review boards of the Cedars-Sinai Medical Center, Standford Healthcare, and Chang Gung Memorial Hospital.

## Reference

1. Anon. How common is hyperkalaemia? A systematic review and meta-analysis of the prevalence and incidence of hyperkalaemia reported in observational studies Toby Humphrey, Mogamat Razeen Davids, Mogamat-Yazied Chothia, Roberto Pecoits-Filho, Carol Pollock. Glen James

2. Anon. Serum Potassium Levels and Mortality in Hemodialysis Patients: A Retrospective Cohort Study Topic Article Package: Topic Article Package: Diabetes Subject Area: Nephrology Content Sponsor: Karger OLA Akeem A. Yusuf; Yan Hu; Bhupinder Singh; José A. Menoyo; James. B. Wetmore

3. Huang C-W, Lee M-J, Lee P-T, et al. Low potassium dialysate as a protective factor of sudden cardiac death in hemodialysis patients with hyperkalemia. PLoS One. 2015;10:e0139886.

4. Bansal S, Pergola PE. Current management of hyperkalemia in patients on dialysis. Kidney Int Rep. 2020;5:779–789.

5. Surawicz B. Relationship between electrocardiogram and electrolytes. Am Heart J. 1967;73:814–834.

6. Wrenn KD, Slovis CM, Slovis BS. The ability of physicians to predict hyperkalemia from the ECG. Ann Emerg Med. 1991;20:1229–1232.

7. Hannun AY, Rajpurkar P, Haghpanahi M, et al. Cardiologist-level arrhythmia detection and classification in ambulatory electrocardiograms using a deep neural network. Nat Med. 2019;25:65–69.

8. Hughes JW, Olgin JE, Avram R, et al. Performance of a convolutional neural network and explainability technique for 12-lead electrocardiogram interpretation. JAMA Cardiol. 2021;6:1285–1295.

9. Chiu I-M, Cheng C-Y, Chang P-K, Li C-J, Cheng F-J, Lin C-HR. Utilization of personalized machine-learning to screen for dysglycemia from ambulatory ECG, toward noninvasive blood glucose monitoring. Biosensors (Basel). 2022;13:23.

10. Yuan N, Duffy G, Dhruva SS, et al. Deep learning of electrocardiograms in sinus rhythm from US veterans to predict atrial fibrillation. JAMA Cardiol. 2023. Published onlineOctober 18, 2023. 10.1001/jamacardio.2023.3701.

11. Theurer J, Yuan. Electrocardiographic deep learning for predicting post-procedural mortality: a model development and validation studyD Ouyang. The Lancet Digital Health. 6:e70–e78.

12. Tison GH, Sanchez JM, Ballinger B, et al. Passive detection of atrial fibrillation using a commercially available smartwatch. JAMA Cardiol. 2018;3:409.

13. Attia ZI, Harmon DM, Dugan J, et al. Prospective evaluation of smartwatch-enabled detection of left ventricular dysfunction. Nat Med. 2022;28:2497–2503.

14. Galloway CD, Valys AV, Shreibati JB, et al. Development and validation of a deep-learning model to screen for hyperkalemia from the electrocardiogram. JAMA Cardiol. 2019;4:428–436.

15. Lin C-S, Lin C, Fang W-H, et al. A deep-learning algorithm (ECG12Net) for detecting hypokalemia and hyperkalemia by electrocardiography: algorithm development. JMIR medical informatics. 2020;8:e15931.

16. Chiu I-M, Cheng J-Y, Chen T-Y, et al. Using deep transfer learning to detect hyperkalemia from ambulatory electrocardiogram monitors in intensive care units: Personalized medicine approach. J Med Internet Res. 2022;24:e41163.

17. Kim D, Jeong J, Kim J, et al. Hyperkalemia detection in emergency departments using initial ECGs: A smartphone AI ECG analyzer vs. Board-certified physicians. J Korean Med Sci. 2023;38.

18. Attia ZI, Kapa S, Lopez-Jimenez F, et al. Screening for cardiac contractile dysfunction using an artificial intelligence-enabled electrocardiogram. Nat Med. 2019;25:70–74.

19. Chen M, Wang G, Ding Z, Li J, Yang H. Unsupervised domain adaptation for ECG arrhythmia classification. Annu Int Conf IEEE Eng Med Biol Soc. 2020;2020:304–307.

20. Anon. HKElectrocardiogram. Apple Developer Documentation Accessed March 26, 2024. https://developer.apple.com/documentation/healthkit/hkelectrocardiogram.

21. Einhorn LM, Zhan M, Hsu VD, et al. The frequency of hyperkalemia and its significance in chronic kidney disease. Arch Intern Med. 2009;169:1156–1162.

22. Luo J, Brunelli SM, Jensen DE, Yang A. Association between serum potassium and outcomes in patients with reduced kidney function. Clin J Am Soc Nephrol. 2016;11:90–100.

23. Cordeiro R, Karimian N, Park Y. Hyperglycemia identification using ECG in deep learning era. Sensors (Basel). 2021;21:6263.

24. Kwon J-M, Jung M-S, Kim K-H, et al. Artificial intelligence for detecting electrolyte imbalance using electrocardiography. Ann Noninvasive Electrocardiol. 2021;26:e12839.

25. Kohsaka S, Okami S, Kanda E, Kashihara N, Yajima T. Cardiovascular and renal outcomes associated with hyperkalemia in chronic kidney disease: A hospital-based cohort study. Mayo Clin Proc Innov Qual Outcomes. 2021;5:274–285.

26. Rafique Z, Hoang B, Mesbah H, et al. Hyperkalemia and electrocardiogram manifestations in end-stage renal disease. Int J Environ Res Public Health. 2022;19:16140.

27. Rafique Z, Aceves J, Espina I, Peacock F, Sheikh-Hamad D, Kuo D. Can physicians detect hyperkalemia based on the electrocardiogram? Am J Emerg Med. 2020;38:105–108.

28. Aslam S, Friedman EA, Ifudu O. Electrocardiography is unreliable in detecting potentially lethal hyperkalaemia in haemodialysis patients. Nephrol Dial Transplant. 2002;17:1639–1642.

29. Kim HE, Cosa-Linan A, Santhanam N, Jannesari M, Maros ME, Ganslandt T. Transfer learning for medical image classification: a literature review. BMC Med Imaging. 2022;22:69.

30. Canali S, Schiaffonati V, Aliverti A. Challenges and recommendations for wearable devices in digital health: Data quality, interoperability, health equity, fairness. PLOS Digit Health. 2022;1:e0000104.

31. Gutierrez Maestro E, De Almeida TR, Schaffernicht E, Martinez Mozos Ó. Wearable-based intelligent emotion monitoring in older adults during daily life activities. Appl Sci (Basel). 2023;13:5637.

32. Vijayakumar V, Ummar S, Varghese TJ, Shibu AE. ECG noise classification using deep learning with feature extraction. Signal Image Video Process. 2022;16:2287–2293.

